# The impact of transmission control measures during the first 50 days of the COVID-19 epidemic in China

**DOI:** 10.1101/2020.01.30.20019844

**Authors:** Huaiyu Tian, Yonghong Liu, Yidan Li, Chieh-Hsi Wu, Bin Chen, Moritz U.G. Kraemer, Bingying Li, Jun Cai, Bo Xu, Qiqi Yang, Ben Wang, Peng Yang, Yujun Cui, Yimeng Song, Pai Zheng, Quanyi Wang, Ottar N. Bjornstad, Ruifu Yang, Bryan T. Grenfell, Oliver G. Pybus, Christopher Dye

## Abstract

Respiratory illness caused by a novel coronavirus (COVID-19) appeared in China during December 2019. Attempting to contain infection, China banned travel to and from Wuhan city on 23 January and implemented a national emergency response. Here we evaluate the spread and control of the epidemic based on a unique synthesis of data including case reports, human movement and public health interventions. The Wuhan shutdown slowed the dispersal of infection to other cities by an estimated 2.91 days (95%CI: 2.54-3.29), delaying epidemic growth elsewhere in China. Other cities that implemented control measures pre-emptively reported 33.3% (11.1-44.4%) fewer cases in the first week of their outbreaks (13.0; 7.1-18.8) compared with cities that started control later (20.6; 14.5-26.8). Among interventions investigated here, the most effective were suspending intra-city public transport, closing entertainment venues and banning public gatherings. The national emergency response delayed the growth and limited the size of the COVID-19 epidemic and, by 19 February (day 50), had averted hundreds of thousands of cases across China.

**One sentence summary:** Travel restrictions and the national emergency response delayed the growth and limited the size of the COVID-19 epidemic in China.

## Main text

On 31 December 2019, less than a month before the 2020 Spring Festival holiday, including the Chinese Lunar New Year, a cluster of pneumonia cases caused by an unknown pathogen was reported in Wuhan, a city of 11 million inhabitants and the largest transport hub in Central China. A novel coronavirus (*1, 2*) was identified as the etiological agent (*3, 4*) and human-to-human transmission of the viral disease (COVID-19) has been since confirmed (*5, 6*). Further spatial spread of this disease was of great concern in view of the upcoming Spring Festival (“*chunyun*”) during which there are typically three billion travel movements over the 40-day holiday period, which runs from 15 days before the Spring Festival (Chinese Lunar New Year) to 25 days afterwards (*7*).

As there is currently neither a vaccine nor a specific drug treatment for COVID-19, a range of public health (non-pharmaceutical) interventions has been used to control the outbreak. In an attempt to prevent further dispersal of COVID-19 from its source, all transport was prohibited in and out of Wuhan city from 10:00h on 23 January 2020, followed by the whole of Hubei Province a day later. In terms of the population covered, this appears to be the largest attempted quarantine (movement restriction) event in human history.

On 23 January, China also raised its national public health response to the highest state of emergency ─ Level 1 of 4 levels of severity in the Chinese Emergency System, defined as an “extremely serious incident” (*8*). As part of the national emergency response, and in addition to the Wuhan city travel ban, suspected and confirmed cases have been isolated, public transport by bus and subway rail suspended, schools and entertainment venues have been closed, public gatherings banned, health checks carried out on migrants (“floating population”), travel prohibited in and out of cities, and information widely disseminated. Despite all these measures, the outbreak has continued to spread geographically, within and beyond China, with mounting numbers of cases and deaths.

Although the spatial spread of infectious diseases has been intensively studied (*9-14*), including explicit studies of the role of human movement (*15, 16*), the effectiveness of travel restrictions and social distancing measures in preventing the spread of infection is uncertain. For COVID-19, coronavirus transmission patterns and the impact of interventions are still poorly understood (*6, 7*). We therefore carried out a quantitative analysis of the impact of travel restrictions and transmission control measures during the first 50 days of the COVID-19 epidemic in China, from 31 December 2019 to 19 February 2020 (Fig. 1). This period embraced the 40 days of the Spring Festival holiday, 15 days before the Chinese Lunar New Year on 25 January and 25 days afterwards. The analysis is based on a unique geocoded repository of data on COVID-19 epidemiology, human movement, and public health (non-pharmaceutical) interventions.

**Figure 1.**
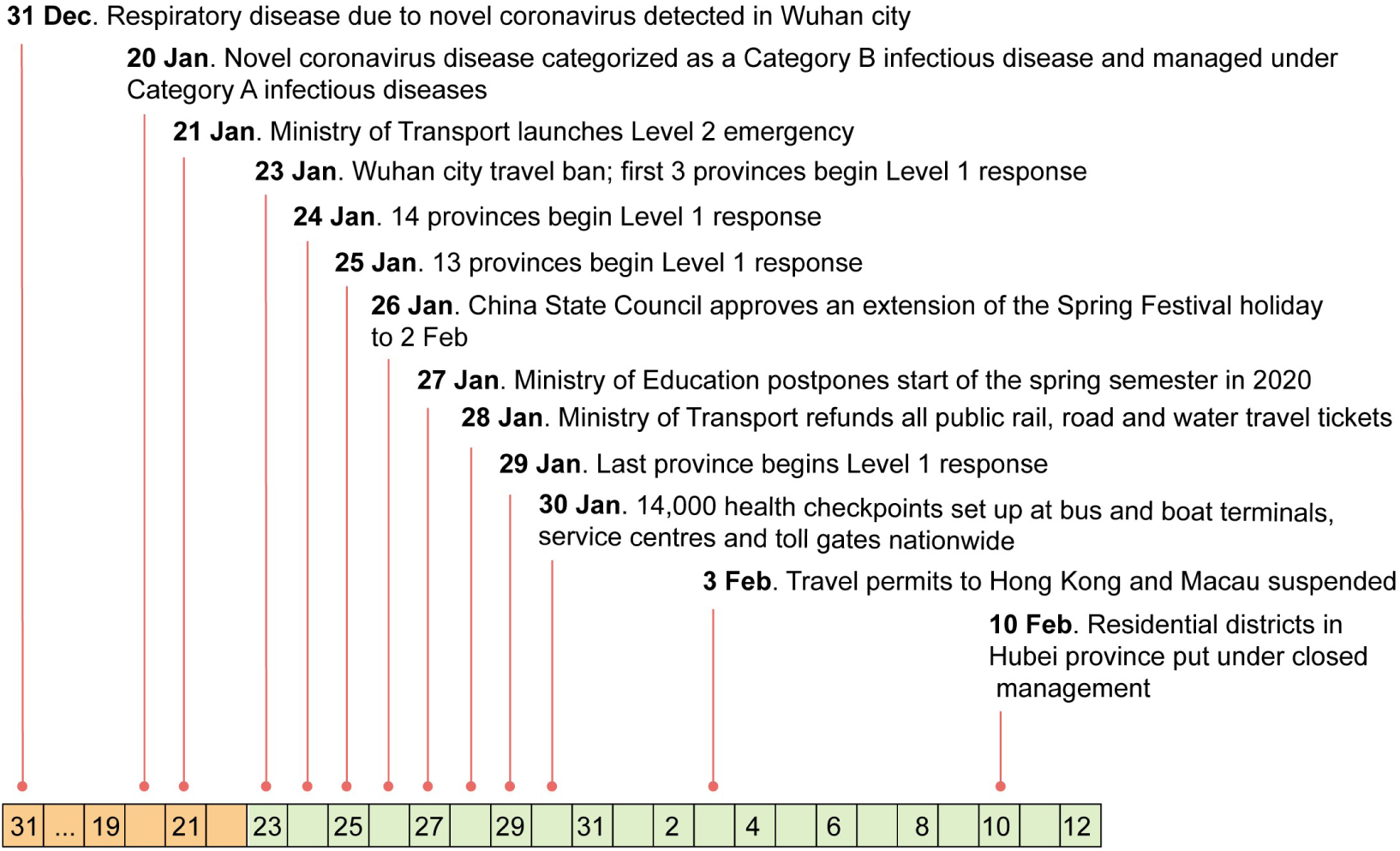
Dates of discovery of the novel coronavirus causing COVID-19, and of the implementation of control measures in China, from 31 December 2019.

We first investigated the effect of the Wuhan city travel ban, comparing travel in 2020 with that in previous years and exploring the consequences of holiday travel for the dispersal of infection across China. During Spring Festival travel in 2017 and 2018, there was an average outflow of 5.2 million people from Wuhan city during the 15 days before the Chinese Lunar New Year. In 2020, this travel was interrupted by the Wuhan city shutdown, but 4.3 million people travelled out of the city between 11 January and the implementation of the ban on 23 January (*7*) (Fig. 2A). In 2017 and 2018, travel out of the city during the 25 days after the Chinese Lunar New Year averaged 6.7 million people each year. In 2020, the travel ban prevented almost all of that movement.

**Figure 2.**
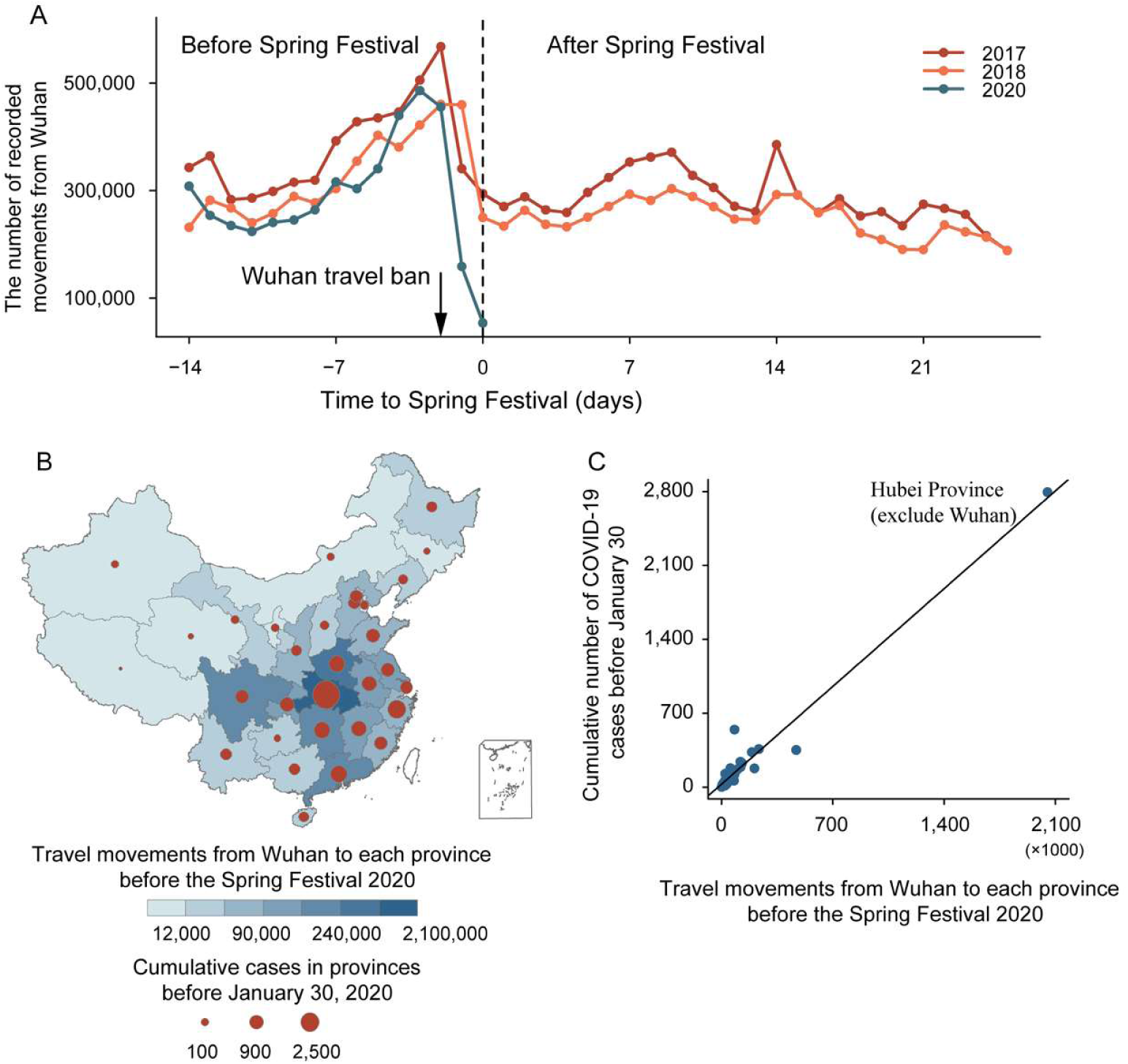
The dispersal of COVID-19 in China 15 days before and 25 days after the Spring Festival (Chinese Lunar New Year). (A) Movement outflows from Wuhan City during Spring Festival travel in 2017, 2018, and 2020. The vertical dotted line is the date of Spring Festival (Chinese Lunar New Year). (B) The number of recorded movements from Wuhan city to other provinces during the 15 days before the Spring Festival in 2020. The shading from light to dark represents the number of human movements from Wuhan to each province. The area of circles represents the cumulative number of cases reported by 30 January 2020, one week after the Wuhan travel ban on 23 January. (C) Association between the cumulative number of confirmed cases reported before 30 January and the number of movements from Wuhan to other provinces.

The dispersal of COVID-19 from Wuhan was rapid (Fig. 3A). A total of 262 cities reported cases within 28 days. For comparison, the 2009 influenza H1N1pdm pandemic took 132 days to reach the same number of cities in China. The number of cities providing first reports of COVID-19 peaked at 59 per day on 23 January, the date of the Wuhan travel ban.

**Figure 3.**
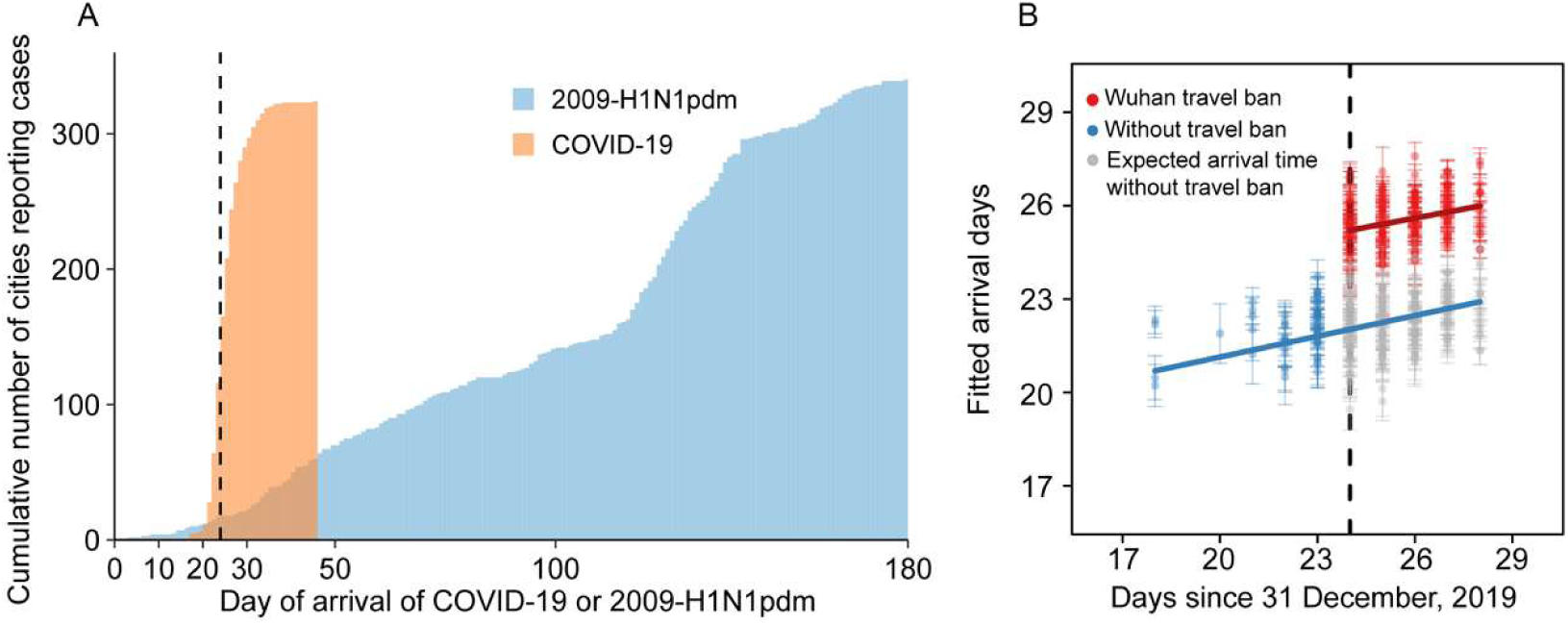
Spatial dispersal of COVID-19 in China. (A) Cumulative number of cities reporting cases by 19 February 2020. Arrival days, defined as the time interval (days) from the date of the first case in the first infected city (Wuhan) to the date of the first case in each newly infected city (a total of 324 cities), to characterize the inter-city transmission rate of COVID-19 and 2009-H1N1pdm, respectively. Dashed line shows the date of Wuhan travel ban (shutdown). (B) Before (blue) and after (red) the intervention by 30 January 2020, one week after Wuhan travel ban (shutdown). The blue line and points show the fitted regression of arrival times up to the shutdown on day 24 (23 January, vertical dashed line). Grey points show the expected arrival times after day 24, without the shutdown. The red line and points show the fitted regression of delayed arrival times after the shutdown on day 24. Each observation (point) represents one city. Error bars give ±2 standard deviations.

The total number of cases reported from each province by 30 January, one week after the Wuhan shutdown, was strongly associated with the total number of travellers from Wuhan (r=0.98, P<0.01; excluding Hubei, r=0.69, P<0.01; Figs. 2B and 2C). COVID-19 arrived sooner in those cities that had larger populations and had more travellers from Wuhan (Tables 1 and S1). However, the Wuhan travel ban delayed the arrival time of COVID-19 in other cities by an estimated 2.91 days (95%CI: 2.54-3.29 days) on average (Table 1, Fig. 3B). More than 130 cities, covering more than half the geographic area and population of China, benefited from the delay.

**Table 1.** Impact of the Wuhan travel ban on COVID-19 dispersal to other cities in China.

| **Covariates** | **Coefficient** | **95% CI** | **P** |
| --- | --- | --- | --- |
| **Intercept** | 25.95 | (23.43, 28.48) | <0.01 |
| **Longitude (degrees)** | -0.03 | (-0.05, -0.01) | <0.01 |
| **Latitude (degrees)** | 0.03 | (0.01, 0.06) | <0.05 |
| **log10 (population)** | -0.70 | (-1.12, -0.28) | <0.01 |
| **log10 (total movements)** | -0.12 | (-0.22, -0.02) | <0.05 |
| **Travel ban (days)** | 2.91 | (2.54, 3.29) | <0.01 |

The dependent variable $Y$ is the arrival time (days) of the outbreak in each city

This delay provided extra time to prepare for the arrival of COVID-19 across China but would not have curbed transmission after infection had been exported to new locations from Wuhan. Fig. 1 shows the timing and implementation of emergency control measures in 342 cities across China (see also Figs. S2 and S4). School closure, the isolation of suspected and confirmed patients, plus the disclosure of information was implemented in all cities. Public gatherings were banned and entertainment venues closed in 220 cities (64.3%). Intra-city public transport was suspended in 136 cities (39.7%) and inter-city travel was prohibited by 219 cities (64.0%). All three measures were applied in 136 cities (Table S2).

Cities that implemented a Level 1 response (any combination of control measures; Figs. S2 and S4) pre-emptively, before discovering any COVID-19 cases, reported 33.3% (95%CI: 11.1-44.4%) fewer laboratory-confirmed cases during the first week of an outbreak (13.0, 95%CI: 7.1-18.8, n=125) compared with cities that started control later (20.6 cases, 95%CI: 14.5-26.8, n=171; difference between groups, U=8197 z=-3.4, *P*<0.01). Among specific control measures, cities that suspended intra-city public transport and/or closed entertainment venues and banned public gatherings, and did so sooner, reported fewer cases during the first week of their outbreaks (Table 2, Table S3). This analysis provided no evidence that the prohibition of travel between cities, which was implemented after and in addition to the Wuhan shutdown on 23 January, reduced the number of cases in other cities across China. These results are robust to the choice of statistical regression model (Supplementary Material, Table S3).

**Table 2.**
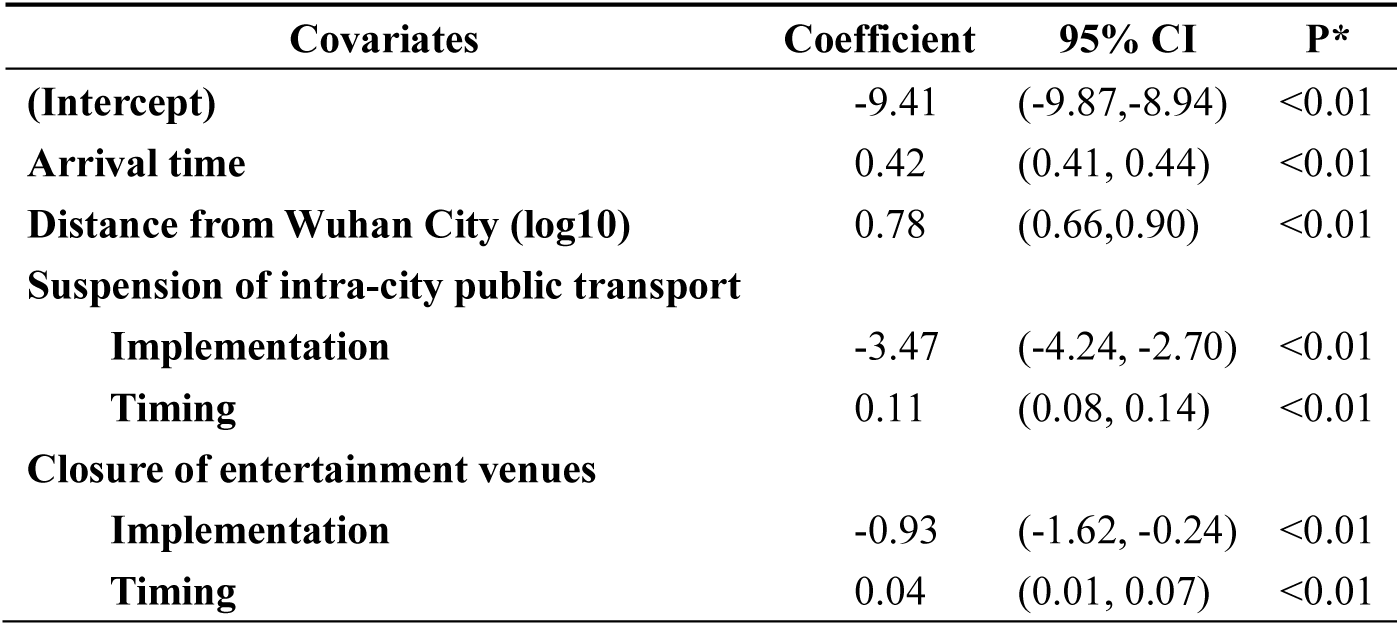
Impact of the type and timing of transmission control measures evaluated by a general linear regression model.

The reported daily incidence of confirmed cases peaked in Hubei province (including Wuhan) on 4 February (3156 laboratory-confirmed cases, 5.33/100,000 population in Hubei), and in all other provinces on 31 January (875 cases, 0.07/100,000 population; Fig. S1). The low level of peak incidence per capita, the early timing of the peak, and the subsequent decline in daily case reports, suggest that transmission control measures not only delayed the growth of the epidemic, but also greatly limited the number of cases. By fitting an epidemic model to the time series of cases reported in each province (Supplementary Material, Fig. S3), we estimate that the (basic) case reproduction number (*R0*) was 3.15 prior to the implementation of the emergency response on 23 January (Table 3). As control was scaled-up from 23 January onwards (stage 1), the case reproduction number declined to 0.97, 2.01 and 3.05 (estimated as *C*_*1*_*R*_*0*_) in three groups of provinces, depending on the rate of implementation in each group (Tables 3 and S4). Once the implementation of interventions was 95% complete everywhere (stage 2), the case reproduction number had fallen to 0.04 on average (*C*_*2*_*R*_*0*_), far below the replacement rate (<< 1) and consistent with the rapid decline in incidence (Fig. 4A, Fig. S5, Table 3, Table S4).

**Table 3.**
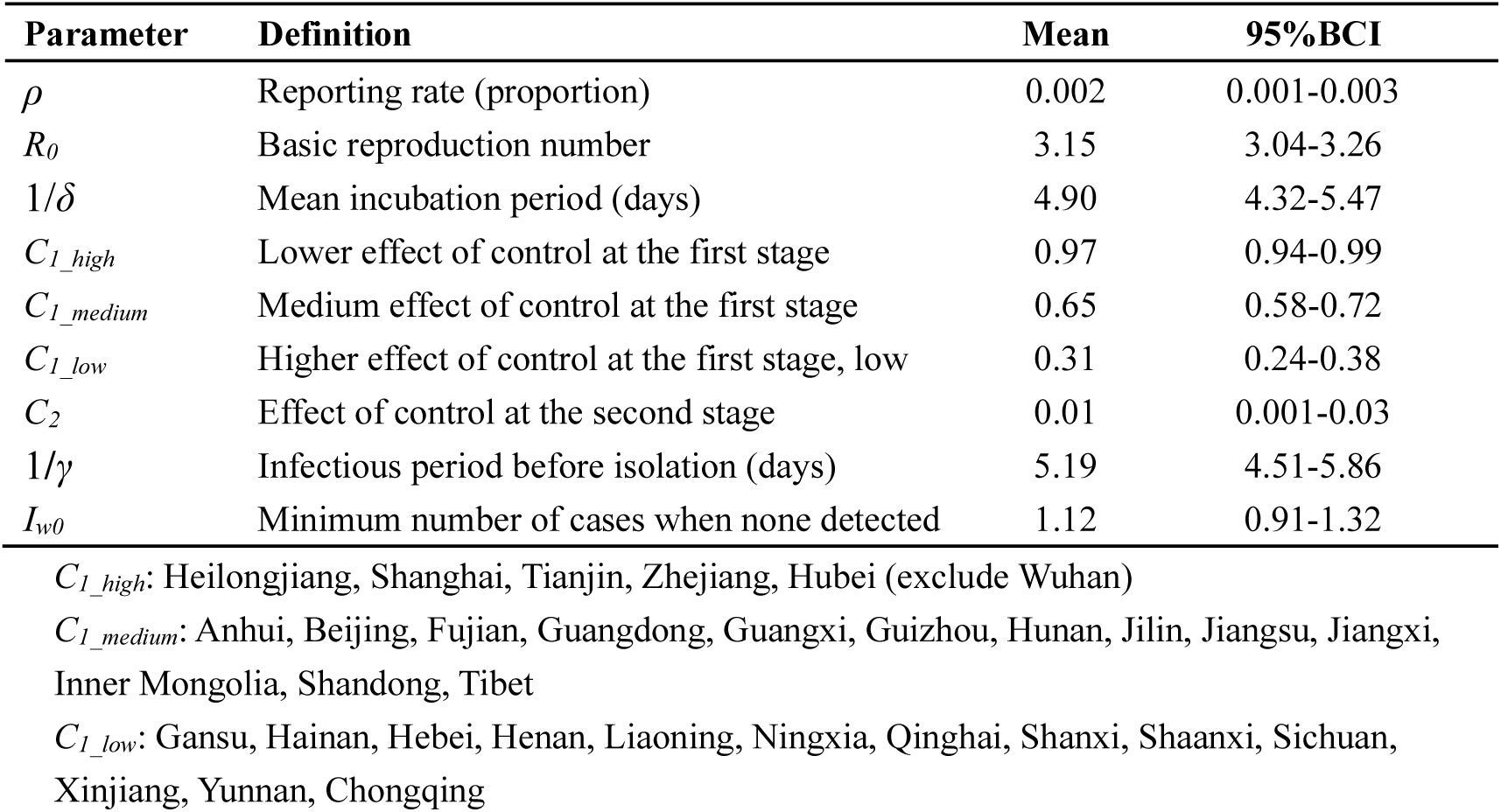
Parameter estimates of the SEIR epidemic model.

**Figure 4.**
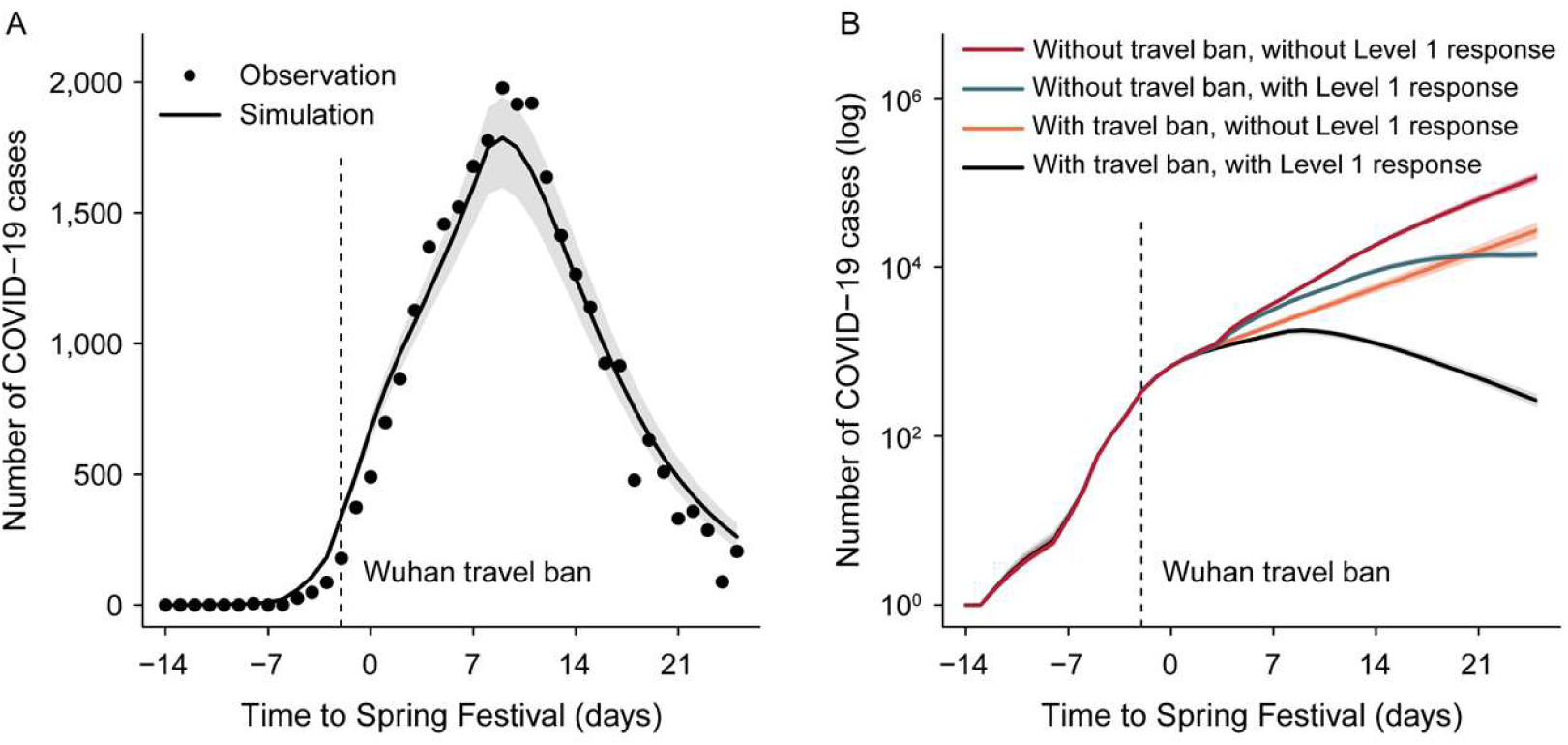
Effect of interventions in controlling the COVID-19 outbreak across China. (A) Epidemic model (line) fitted to daily reports of confirmed cases (points) summed across 31 provinces. Hubei excludes Wuhan city. (B) Expected epidemic trajectories without the Wuhan travel ban (shutdown), and with (green) or without (red) interventions carried out as part of the Level 1 national emergency response; with the Wuhan travel ban, and with (black) or without the intervention (orange). Vertical dashed lines in both panels mark the date of the Wuhan travel ban and the start of the emergency response, on 23 January. Shaded regions mark the 95% Bayesian credible intervals.

Based on the fit of the model to daily case reports from each province, we investigated the aggregate effect of control measures on the trajectory of the epidemic outside Wuhan city (Fig. 4B). Without the Wuhan travel ban or the national emergency response, there would have been 744,000 (± 156,000) confirmed COVID-19 cases outside Wuhan by 19 February, day 50 of the epidemic. The Wuhan travel ban alone would have reduced this number to 202,000 (± 10,000) cases, by delaying epidemic growth. The national emergency response alone would have cut the number of cases to 199,000 (± 8500). Therefore neither of these interventions would, on their own, have reversed the rise in incidence by 19 February (Fig. 4B). But together and interactively, these control measures evidently did halt and reverse the rise in incidence, limiting the number of confirmed cases reported to 29,839 (fitted model estimate 28,000 ± 1400 cases), a 96% reduction on the total number of cases expected in the absence of interventions.

In summary, this early analysis suggests that transmission control (non-pharmaceutical) measures initiated during Chinese Spring Festival holiday, including the unprecedented Wuhan city travel ban and the Level 1 national emergency response, delayed the growth and limited the size of the COVID-19 epidemic in China. Urbanization and the development of rapid transport systems in China (*17-20*) probably accelerated the spread and magnified the challenge of controlling COVID-19, as indicated by the comparatively slow dispersal of pandemic influenza H1N1pdm in 2009. In addition, the COVID-19 epidemic began just before the period of intense travel during the Spring Festival holiday. Nevertheless, the Wuhan city travel ban provided extra time to implement transmission control measures in other parts of China and, once established, these were an additional powerful force in curtailing and reversing the epidemic.

The number of people who have developed COVID-19 during this epidemic, and therefore the number of people who were protected by control measures, is not known precisely, given that cases were almost certainly under-reported. However, in view of the small fraction of people known to have been infected by 19 February (75,532 cases, 5.41 per 100,000 population), it is unlikely that the spread of infection was halted and epidemic growth reversed because the supply of susceptible people had been exhausted. This implies that a large fraction of the Chinese population remains at risk of COVID-19; relaxing control measures could lead to a resurgence of transmission. Further investigations are needed to verify that proposition, and population surveys of infection are needed to reveal the true number of people who have been exposed to this novel coronavirus.

We could not investigate the impact of all elements of the national emergency response because many were introduced simultaneously across China. However, there is firm evidence from the data used in this analysis that suspending intra-city public transport, closing entertainment venues and banning public gatherings, which were introduced at different times in different places, contributed to the overall containment of the epidemic. Other factors are likely to have contributed to control, such as the isolation of suspected and confirmed patients, contact tracing and the closure of schools, and it is not yet clear which parts of the national emergency response were most effective. We did not find that prohibiting travel between cities or provinces reduced the numbers of COVID-19 cases outside Wuhan and Hubei, perhaps because such travel bans were implemented as a response to, rather than in anticipation of, the arrival of COVID-19.

This study has drawn inferences, not from a controlled experiment, but from statistical and mathematical analyses of the temporal and spatial variation in case reports, human mobility and transmission control measures. With that caveat, we conclude that these control measures had a major impact on the COVID-19 epidemic, averting hundreds of thousands of cases by 19 Feburary. Whether the means and the outcomes of control can be replicated outside China, and which of the interventions are most effective, are now under intense investigation as the virus continues to spread worldwide.

## Data Availability

Population sizes for each city were collected from the China City Statistical Yearbook (http://olap.epsnet.com.cn/).Human movement can be observed directly from mobile phone data, through the location-based services (LBS) employed by popular Tencent applications, such as WeChat and QQ. Average movement outflows from Wuhan City to other cities, by air, train, and road, were calculated from the migration flows database (https://heat.qq.com/) over the entire 2018.

## Acknowledgements

We thank the thousands of CDC staff and local health workers in China who collected data and continue to work to contain COVID-19 in China and elsewhere. Funding for this study was provided by the Beijing Natural Science Foundation (JQ18025); Beijing Advanced Innovation Program for Land Surface Science; National Natural Science Foundation of China (81673234); Young Elite Scientist Sponsorship Program by CAST (YESS)(2018QNRC001); HT, OGP and CD acknowledge support from the Oxford Martin School; HT acknowledges support from the Military Logistics Research Program. The funders had no role in study design, data collection and analysis, the decision to publish, or in preparation of the manuscript.

## Supplementary Material

### Materials and Methods

#### Data sources

##### Epidemiological, demographic and geographical data

We collected data from the official reports of the health commission of 34 provincial-level administrative units and 342 city-level units. We recorded the date of the first reported case in all newly-infected cities, including daily reports from 31 December 2019 to 19 February 2020, the first 50 days of the epidemic. Only laboratory-confirmed cases of COVID-19 were used.

Population sizes for each city in 2018 were obtained from the China City Statistical Yearbook (http://olap.epsnet.com.cn/). Using ArcGIS we calculated the great circle distance between Wuhan and each city reporting COVID-19 cases. The location of each city is geocoded by the latitude and longitude coordinates of the city centre. For the 2009 H1N1 Pandemic (2009-H1N1pdm), daily case data were collected from China Information System for Disease Control and Prevention (CISDCP) from 10 May 2009 to 30 April 2010, a total of more than 180,000 cases (*5*).

##### Human mobility data

Human movements were tracked with mobile phone data, through location-based services (LBS) employed by popular Tencent applications such as WeChat and QQ. Movement outflows from Wuhan City to other cities (i.e. records of the number of people leaving each day) by air, train and road, were obtained from the migration flows database (https://heat.qq.com/) (*21*) from 13 January 2017 to 21 February 2017 (Spring Festival travel 2017), from 1 February 2018 to 12 March 2018 (Spring Festival travel 2018), and from 1 January 2018 to 31 December 2018 (entire 2018). To reconstruct the movement outflow from Wuhan during the 2020 Spring Festival (from 11 January to 25 January, before the Chinese Lunar New Year), mobile phone data (provided by the telecommunications operators) were used together with the Baidu migration index (http://qianxi.baidu.com/); using both data sources gave the most accurate measure of movement volume. The expected movement outflows from Wuhan after the New Year festival from 26 January to 19 February, had there been no travel ban, were generated by using travel volumes for 2017 and 2018 and the recorded travel destinations prior to the shutdown in 2020. We assumed that the proportion of daily outflows from Wuhan to each of the other destinations in China was constant through time.

#### Data analysis

##### Effect of the Wuhan city travel ban on the arrival time of COVID-19 in other cities

In order to quantify the effect of the Wuhan travel shutdown (23 January 2020) on COVID-19 spread, we used data collected between 31 December 2019 and 28 January 2020. The association between distance, human movement, interventions and timing of COVID-19 spread was assessed by regression with a general linear model (GLM). Among five possible regression models examined (Table S3), the model judged best by the Akaike Information Criterion) was:

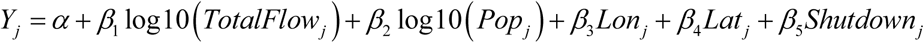

Dependent variable *Y*_*j*_ is the arrival time (day) of the first confirmed case in city *j*, a measure of the spatial spread of COVID-19. The *β*_*i*_ are the regression coefficients. *α* is the intercept. *TotalFlow*_*j*_ represents the passenger volume from Wuhan to city *j* by airplane, train and road during the whole of 2018. *Pop*_*j*_ is the population of city *j. Lat*_*j*_ and *Lon* _*j*_ represent the latitude and longitude of city *j*. The binary dummy variable *Shutdown*_*j*_ is used to identify whether the arrival time of COVID-19 in newly-infected city *j* is influenced by the Wuhan travel ban. For each city, *shutdown* was set to 0 for arrival before 23 January 2020 and 1 for arrival on or after 23 January 2020. The regression analysis was performed using the R package (R version 3.4.0) MASS (*22*). All of the candidate models examined (Table S3) produced similar estimates for the estimated delay in the arrival time due to the shutdown.

##### Effect of transmission control measures on the number of cases reported during the first week of an outbreak in a new location

The Level 1 national emergency response required suspected and confirmed cases of COVID-19 to be isolated and reported immediately in all cities. Using data for 342 cities across China, we investigated the effects of three transmission control measures: closure of entertainment venues and banning public gatherings (*B*); suspension of intra-city public transport (*S*); and prohibition of travel by any means to and from other cities (*P*). The timing of implementation was recorded for each control measure in each city, including the delay in implementation since 31 December 2019 (day 0 of the epidemic). Each city was regarded as implementing an intervention when the official policy was announced publicly (Table S1). Other transmission control measures included delineating control areas, closure of schools, isolation of suspected and confirmed cases, and the disclosure of information. The effects of these interventions could not be investigated because they were reportedly applied in all cities uniformly and without delay.

As above, we used regression analysis to investigate the effects of interventions *B, S* and *P*. The dependent (Poisson) variable is the total number of confirmed cases that were reported during the first seven days (*μ*) of an outbreak in any city (*i*). The analysis was performed using the GLM function in the statistical software R (version 3.6.2) using the model:

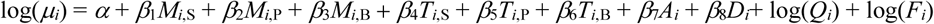

here population size of a city *i* (*O*_*i*_) and inflow from Wuhan (*F*_*i*_) are offset variables, while the distance to Wuhan and the arrival time of the infection are adjustments to control for confounding with other independent variables. The *β*_*j*_’s are regression coefficients. *M*_*i*_,_*k*_ is a binary variable indicating whether or not control measure *k* is implemented in city *i. T*_*i,k*_ represents the timing of implementation of control measure *k* in city *i. D*_*i*_ is the distance from city *i* to Wuhan City. *A*_*i*_ is the arrival time of the epidemic in city *i* (the date of the first confirmed case).

To check and confirm the validity of results obtained with the Poisson regression model, we repeated the analysis with a log-linear model. The first step was to standardize case counts by dividing by the number of people in each city (incidence per capita) and the number of people arriving from Wuhan, giving dependent variable *ν*. The log-linear model is then:

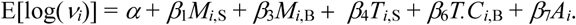

The subscripts of the coefficients (*β*_*j*_) are consistent with a Poisson regression model. To avoid heteroscedasticity, variables describing the distance from Wuhan, and the implementation and timing of *P* (prohibiting inter-city travel) were removed. Further exploration of the model showed that these variables did not help to explain variation in *ν*. Table S3 presents the results of the log-linear regression analysis, which uphold the conclusions reached from the Poisson regression model.

##### Epidemic modelling

For each province, we estimated the effect of transmission control measures by fitting an SEIR model (*23*) to the number of new confirmed cases reported each day from each province using Bayesian Markov Chain Monte Carlo methods (*24*). The model is:

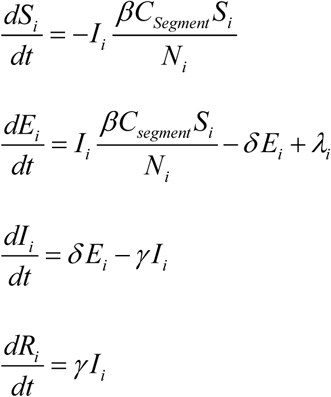

here *S, E, I*, and *R* are the number of susceptible, exposed (latent), infectious, and removed individuals on day *t* in province *i*. This standard SEIR model makes some simplifying assumptions: for example, the human population is homogeneous (e.g. not stratified by age or sex), contacts between infectious and susceptible people are also homogeneous (e.g. not stratified by social group) and infection is fully immunizing (*1*). However, the model describes the data accurately (Fig. 4A, Fig. S3) and these assumptions are unlikely to affect the principal conclusions of the analysis, which apply only to the first 50 days of the epidemic. The basic reproductive number of the model is *R*_*0*_ =*β*/*γ*, where *β* is the per capita transmission rate per day and 1/*δ* and 1/*γ* are, respectively, the mean latent and infectious periods.

Variable *λ* is the estimated number of cases imported from Wuhan City on day *t*:

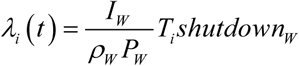

*I*_*w*_ is the number of reported cases in Wuhan on day *t, P*_*w*_ is the Wuhan population size, and *ρ*_*w*_ is the proportion of all infected people (including infectious cases) reported in Wuhan. *T*_*i*_ is the number of people leaving Wuhan on day *t* travelling to province *i*, derived from data describing mobility 15 days before the Chinese Lunar New Year 2020. The binary variable *shutdown* is used to identify whether cases were or were not exported from Wuhan on or after 23 January 2020.

The effects of control measures at different stages of the outbreak are captured by estimated parameter *C* (range 0-1), which reduces transmission and *R*_*0*_ proportionally as a multiplicand of *β*. The timing and implementation of transmission control measures in the 342 cities and 31 provinces are shown in Fig. S4. Before 22 January 2020, there were no recorded interventions thus *C*_*0*_=1. From 23 January onwards, provinces gradually scaled up Level 1 emergency responses (stage 1), with effects measured as *C*_*1*_ (Fig. S4). Because the effects of control measures varied among provinces during the scale-up, *C*_*1*_ was grouped into high *C*_*1_high*_, medium *C*_*1_medium*_, and low C_*1_low*_. The allocation of provinces to groups was made by proposing several alternative hypotheses and testing each by model fitting (Table 3, Table S4). Stage 2 of control (*C*_*2*_) began when more than 95% of cities in a province had implemented control measures, including the closure of entertainment venues, suspension of intra-city public transport or prohibition of travel by any means to and from other cities (see above). In Hubei Province (except Wuhan city), stage 2 included the use of shelter or “Fang Cang” hospitals from early February onwards.

Model fitting was performed using the Metropolis–Hastings Markov chain Monte Carlo (MCMC) algorithm with the MATLAB (version R2016b) toolbox DRAM (Delayed Rejection Adaptive Metropolis). Prior estimates of the mean and (Gaussian) variance of *R*_*0*_, *δ*, and *γ* were derived from epidemiological surveys (*25*). There was no evidence to inform a prior for the reporting rate *ρ*, the proportion of cases that were reported among all latent and infectious individuals in Wuhan.

Systematic surveys of infection (e.g. by serological testing) have not yet been reported. In the absence of any guiding data, *ρ* was given a prior uniform distribution between 0 and 1.

After a burn-in of 1 million iterations, we ran the MCMC simulation for a further 10 million iterations, sampled at every 1000th step to avoid auto-correlation. Trace plots and Gelman and Rubin diagnostics were used to judge convergence of the MCMC chains (Fig. S4). Each fitting exercise was repeated three times to test the robustness of results, which converged to the same estimates on each occasion (Fig. S5). We used the fitted SEIR model, with posterior estimates of parameter values, to simulate outbreaks outside Wuhan, with and without the Wuhan travel ban and with and without the national emergency response (Fig. 4B).

## Supplementary Figures and Tables

**Figure S1.**
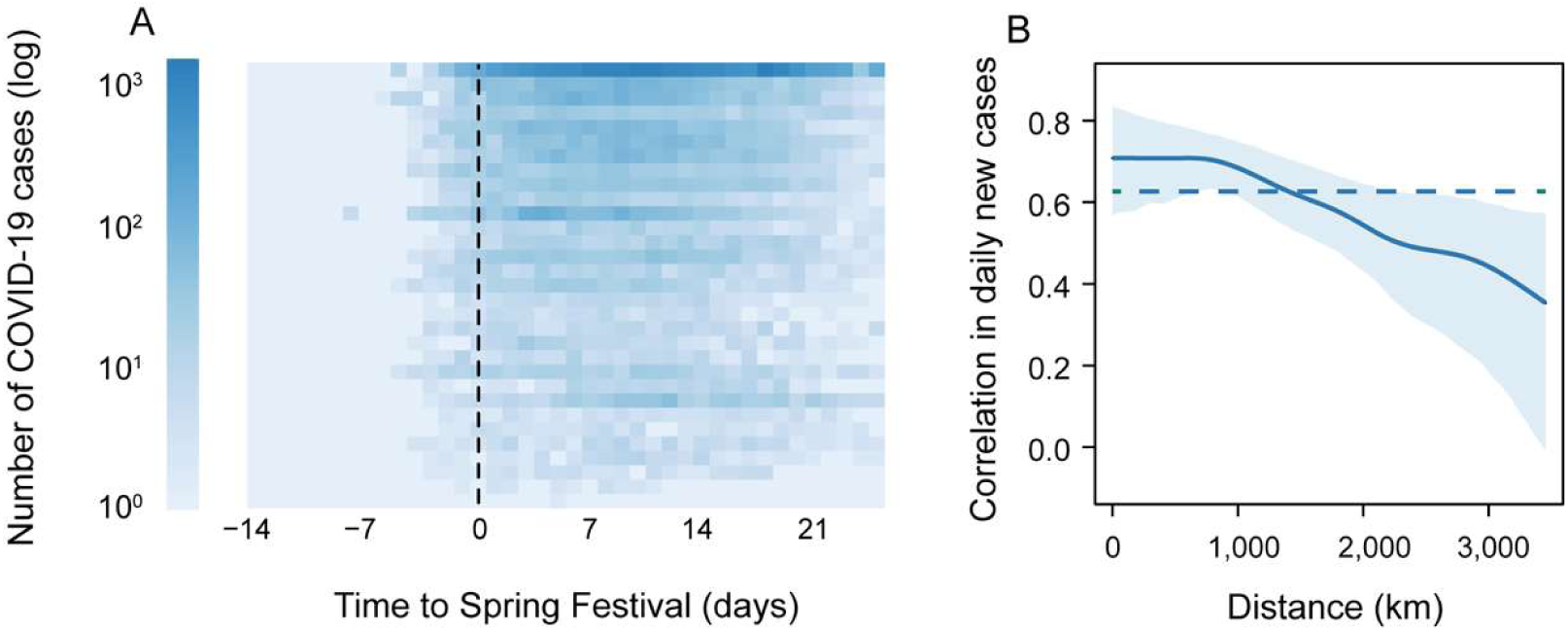
Patterns of COVID-19 dispersal out of Wuhan (Hubei province) to other provinces by time and geographical distance. (A) Daily reports of confirmed cases from each of 31 provinces. Provinces are ranked by decreasing volume of people leaving Wuhan for other destinations, to elsewhere in Hubei Province (top) and to Tibet (bottom). (B) Synchrony of epidemics in different provinces in relation to distance between provinces. Synchrony is measured by the correlation between the number of cases reported in two provinces on each day, using a spatial non-parametric correlation function (*26*).

**Figure S2.**
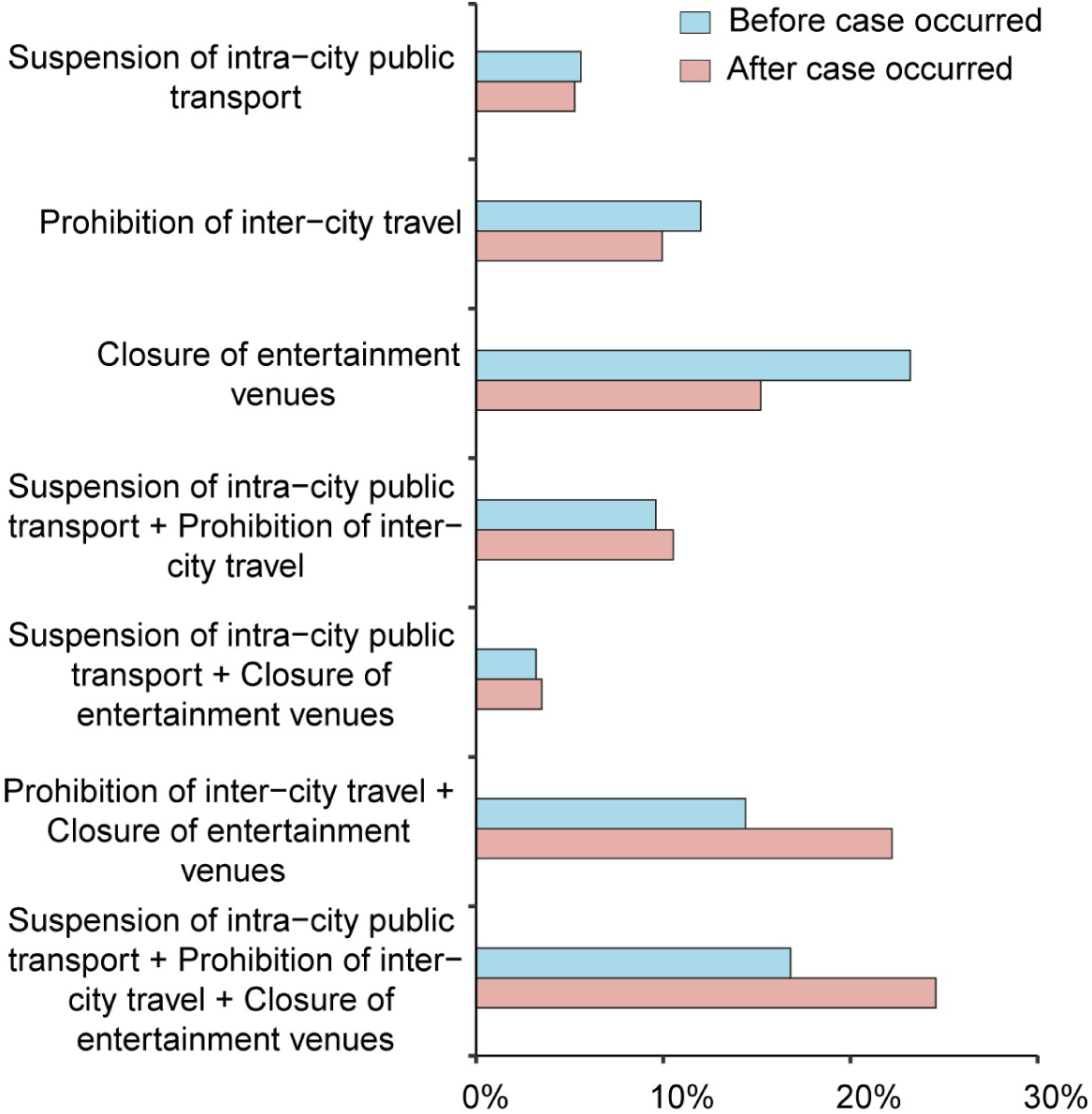
Percentage of cities that implemented three kinds of transmission control measures before (blue), or on the same day or after (red), the first case was reported.

**Figure S3.**
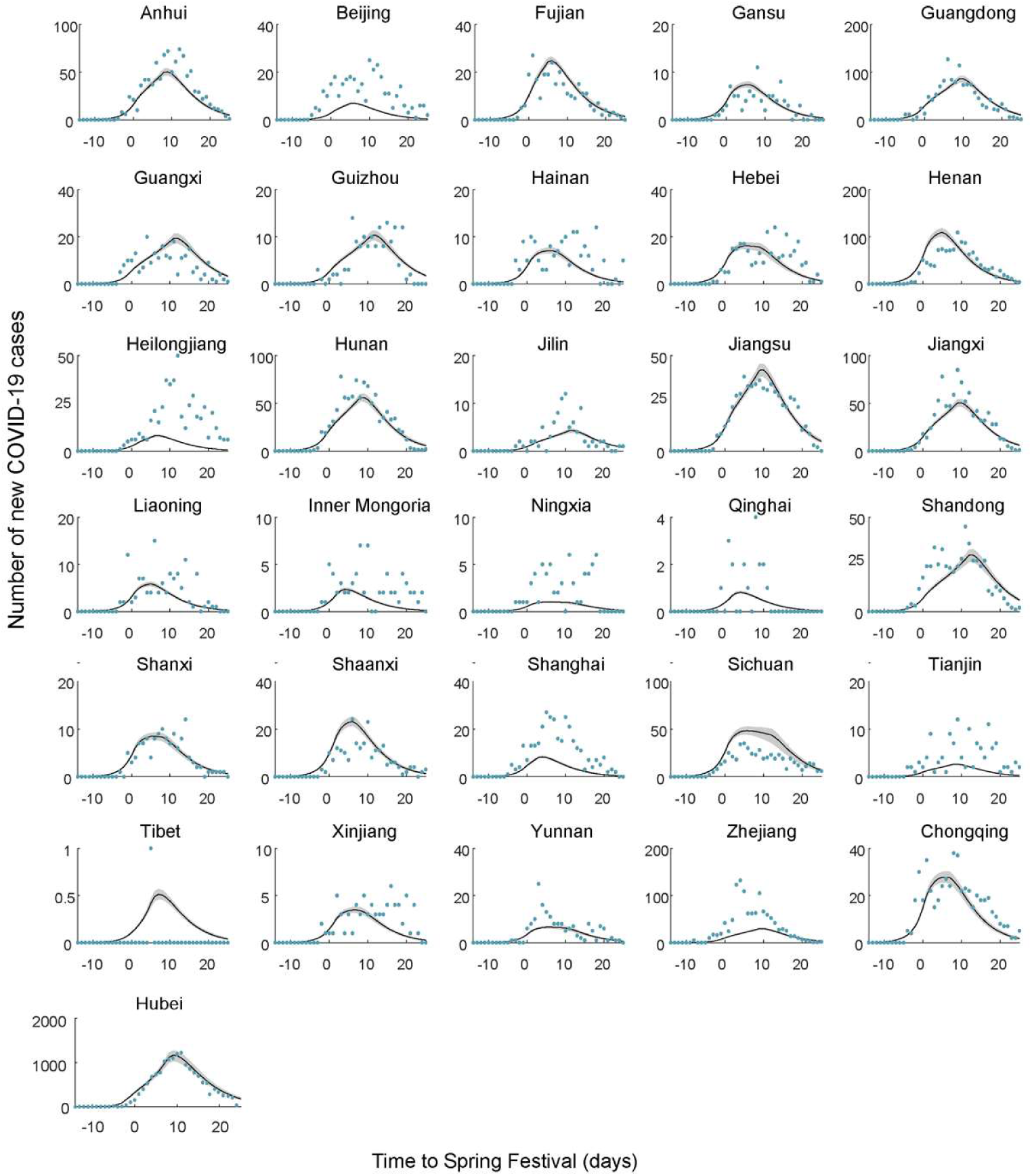
Fits of the SEIR epidemic model to time series of reported cases from 31 provinces. The numbers of confirmed cases reported (points) and estimated (lines) each day in each provinces (Hubei excludes Wuhan city). Grey areas correspond to pointwise 95% credible envelopes. The period covers the 40 days of the Spring Festival, from 15 days before to 25 days after the Chinese Lunar New Year. The Spring Festival holiday ended on 19 February, day 50 of the epidemic.

**Figure S4.**
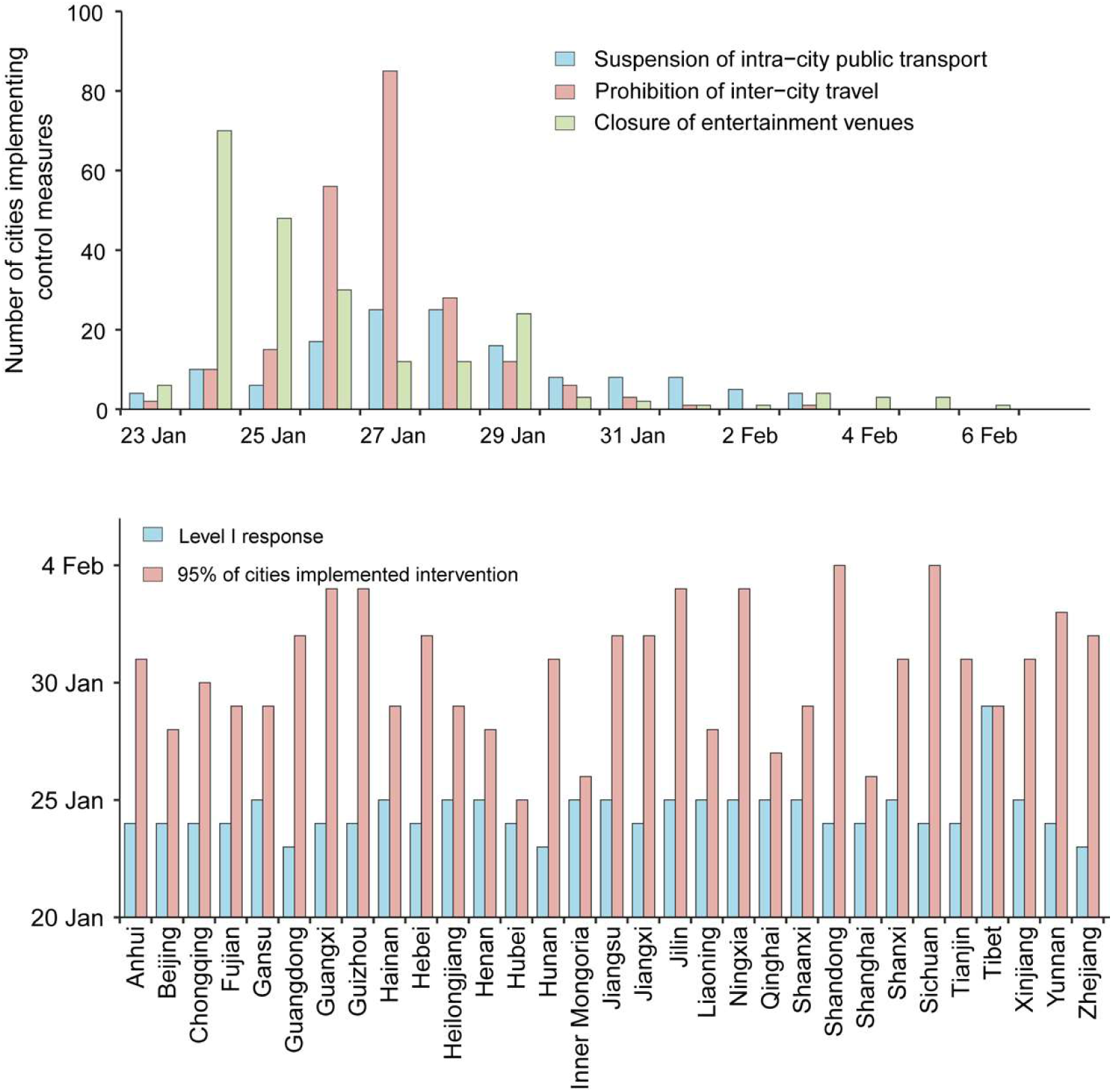
(A) The number of cities implementing three interventions by date in 342 cities (see also Fig. S2). (B) Dates (vertical axis) on which the Level 1 emergency response began (blue, start of stage 1), and on which 95% of cities had implemented transmission control measures (red, end of stage 1, beginning of stage 2), in 31 provinces.

**Figure S5.**
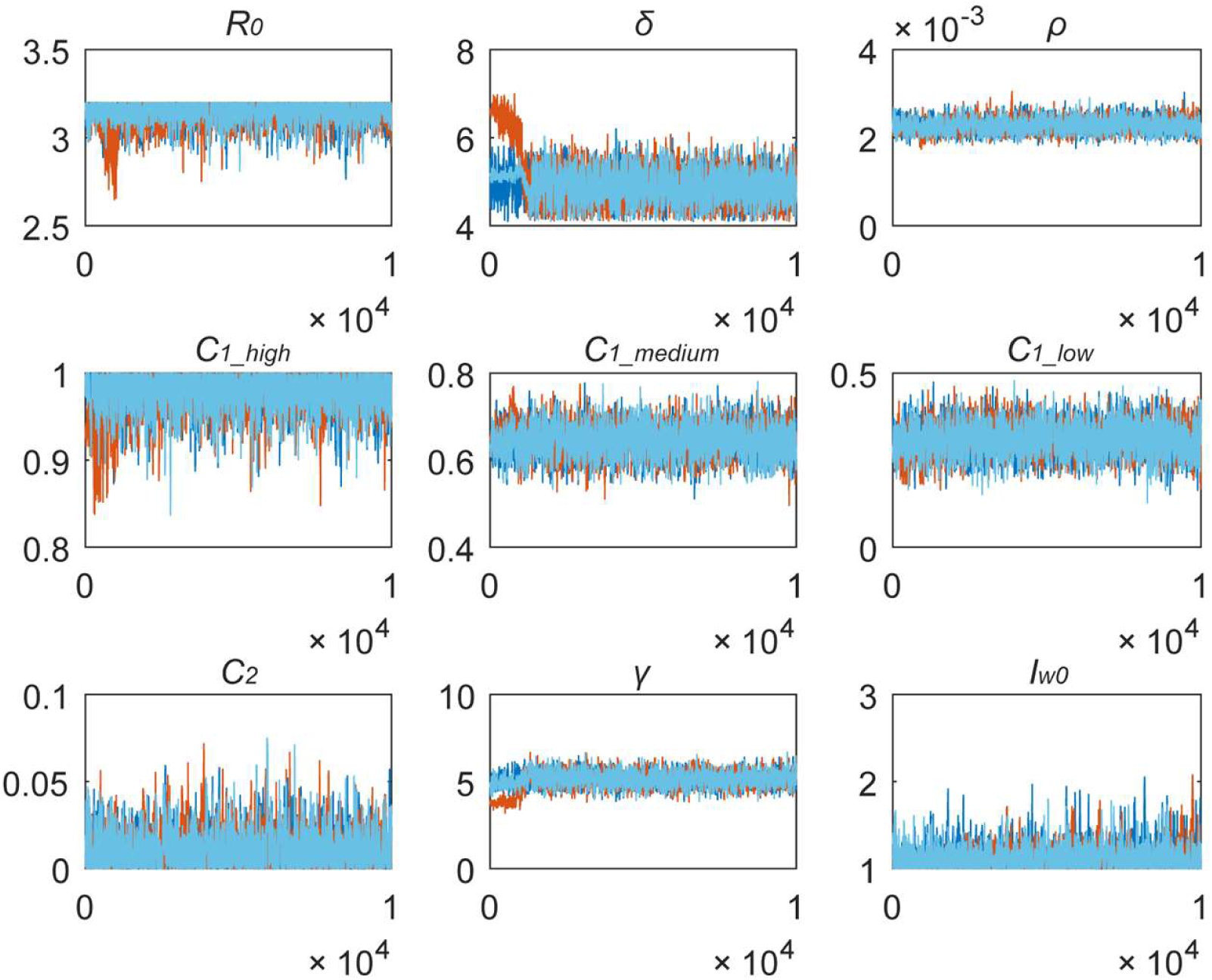
Trace plots of parameter values for the epidemic model, estimated by Bayesian Markov Chain Monte Carlo (MCMC) methods. The three different colours represent three runs of the MCMC model, with one run (light blue) presented at the forefront.

**Table S1.** Candidate statistical models used to study the effect of the Wuhan city travel ban on the arrival time of COVID-19 in other cities (see Table 1 of the main text).

| Model | AIC* |
| --- | --- |
| **Y=log10(TotalFlow) + log10(AirFlow) + log10(RoadFlow) + log10(TrainFlow) + log10(Pop) + log10(Dis) + lat + lon + Shutdown** | 113.73 |
| **Y=log10(TotalFlow) + log10(AirFlow) + log10(RoadFlow) + log10(Pop) + log10(Dis) + lat + lon + Shutdown** | 111.91 |
| **Y=log10(TotalFlow) + log10(AirFlow) + log10(Pop) + log10(Dis) + lat + lon + Shutdown** | 110.63 |
| **Y=log10(TotalFlow) + log10(Pop) + log10(Dis) + lat + lon + Shutdown** | 109.83 |
| **Y=log10(TotalFlow) + log10(Pop) + lat + lon + Shutdown** | 108.57 |

\* Akaike information criterion

**Table S2.** Summary of interventions and their timing across 342 cities (see Table 2 of the main text).

| **Level 1 response to major public health emergencies** | **Number of cities**<br>**implementing**<br>**control measures** | **Average lags (days)**<br>**between implementation**<br>**and 31 December 2019‡** |
| --- | --- | --- |
| **Identify the affected area of a city\*** | 342 | 0 |
| **Close schools\*** | 342 | 0 |
| **Close entertainment venues and ban public gatherings** | 220 | 27.17 (2.82) |
| **Isolate patients with infectious diseases\*** | 342 | 0 |
| **Isolate suspected patients\*** | 342 | 0 |
| **Suspend intra-city public transport (bus and subway)** | 136 | 29.00 (2.60) |
| **Prohibit inter-city travel** | 219 | 27.86 (1.49) |
| **Collect, evaluate, report and publish information on**<br>**public health emergencies daily\*** | 342 | 0 |
| **Assist subdistrict, township (town), neighbourhood and**<br>**village committee staff\*** | 342 | 25.32 (1.07) |

\*Interventions implemented immediately were not included in the regression analysis.
‡Summary statistics reported for timing are mean (standard deviation).

**Table S3.** Impact of the type and timing of transmission control measures, estimated from a log-linear regression model. This analysis checks and confirms the robustness of results in Table 2 of the main text. As described in the main text, the prohibition of inter-city travel, the third intervention that was investigated in this study, did not significantly reduce the number of cases reported during the first week of city outbreaks.

| Covariates | Coefficient | 95% CI | P |
| --- | --- | --- | --- |
| **(Intercept)** | -1.69 | (-5.64, 2.26) | 0.40 |
| **Arrival time** | 0.28 | (0.13, 0.43) | <0.01 |
| **Suspension of intra-city public transport** |  |  |  |
| **Implementation** | -12.65 | (-21.71, -3.60) | <0.01 |
| **Timing** | 0.46 | (0.13, 0.79) | <0.01 |
| **Closure of entertainment venues** |  |  |  |
| **Implementation** | -3.44 | (-5.75, -1.13) | <0.01 |
| **Timing** | 1.53 | (0.69, 2.38) | <0.01 |

**Table S4.**
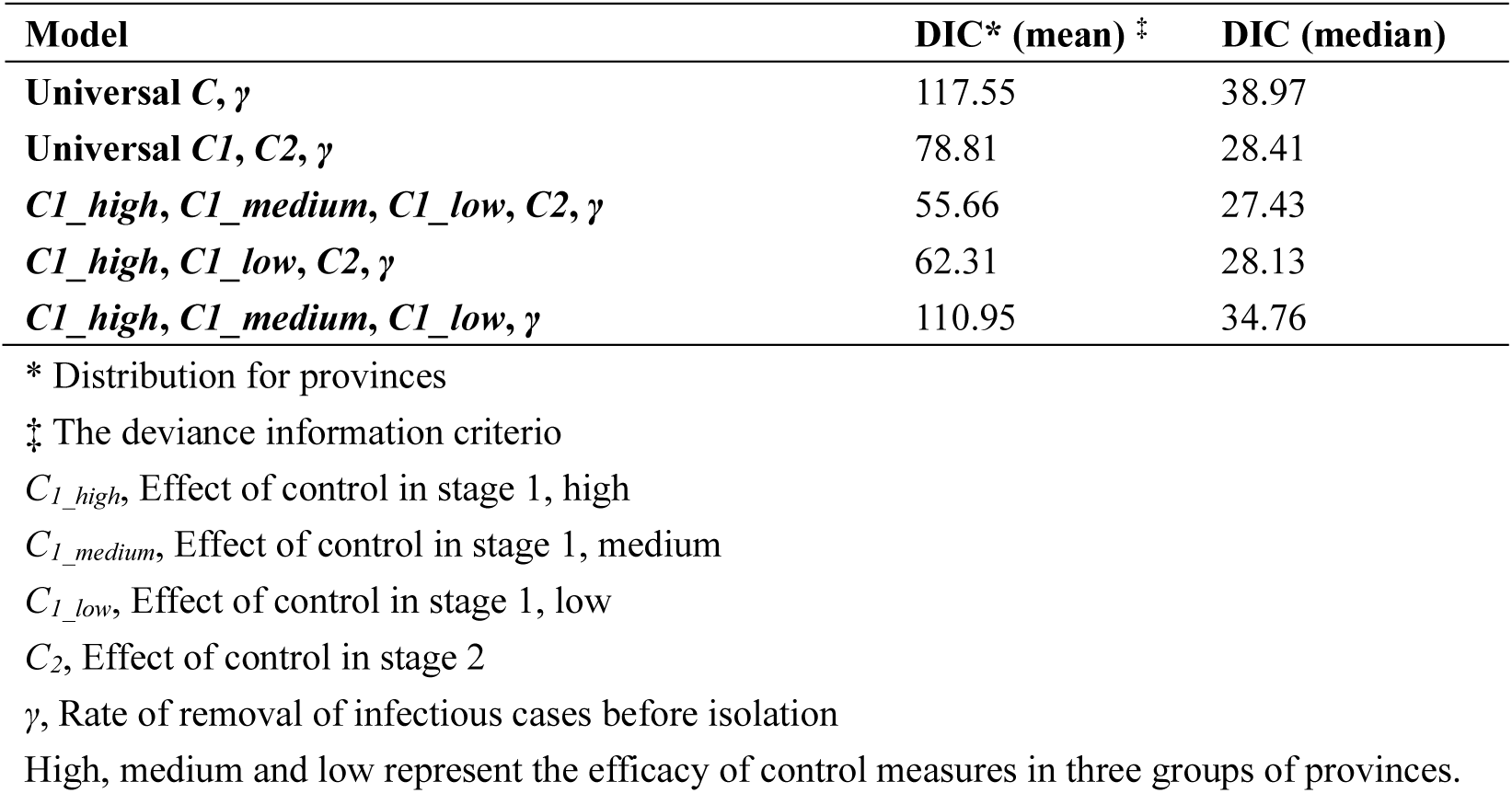
Candidate models used to characterize the effect of control measures in different provinces (see Table 3 of the main text).

| **Model** | **DIC\* (mean) ‡** | **DIC (median)** |
| --- | --- | --- |
| **Universal $C, \gamma$** | 117.55 | 38.97 |
| **Universal $C1, C2, \gamma$** | 78.81 | 28.41 |
| **$C1_{high}, C1_{medium}, C1_{low}, C2, \gamma$** | 55.66 | 27.43 |
| **$C1_{high}, C1_{low}, C2, \gamma$** | 62.31 | 28.13 |
| **$C1_{high}, C1_{medium}, C1_{low}, \gamma$** | 110.95 | 34.76 |

\* Distribution for provinces
‡ The deviance information criterio
$C1_{high}$ , Effect of control in stage 1, high
$C1_{medium}$ , Effect of control in stage 1, medium
$C1_{low}$ , Effect of control in stage 1, low
$C2$ , Effect of control in stage 2
$\gamma$ , Rate of removal of infectious cases before isolation
High, medium and low represent the efficacy of control measures in three groups of provinces.

